# What is the role and value of facilitation within sessions of Case-based Learning (CBL) in undergraduate medicine: Protocol for a scoping review of the literature

**DOI:** 10.1101/2021.11.04.21265716

**Authors:** C. Berg, L. Hammond, B. Howard, B. Diug, O. Sorinola

## Abstract

**Background:** CBL involves students working within groups to solve clinical and non-clinical cases which enables the development of communication and team-working skills and promotes a deeper understanding of relevant clinical knowledge. CBL sessions are traditionally supported by an in-person facilitator in face to face settings. The recent COVID-19 pandemic has necessitated moves to online learning in many areas, one of which is CBL, providing good reason to consider the role of the facilitator in CBL and digital opportunities to augment facilitation.

**Aims:** To understand the role of facilitation within CBL teaching (in face to face and online settings) and the impact that different approaches to facilitation can have on outcomes including student experience and engagement.

**Methods:** A scoping review will be conducted of four main academic and medical databases: MedLine, Scopus, Excerpta Medica Database (EMBASE) and Education Research Complete (ERC) using a search strategy built around ‘Undergraduate medical students’ (Population), ‘Approaches to facilitation’ (Concept) and ‘Case-based learning’ (Context). Articles will be selected according to pre-determined inclusion and exclusion criteria, with scope for the selection criteria to further develop. Charting from included full text articles will encompass research approach, approach to facilitation and findings about facilitation. A constructivist conceptual framework will be used to guide the synthesis.

**Conclusions:** Overall, it is hoped that this review will be able to inform the role of facilitation within CBL sessions and the potential impact that this has on attainment, engagement and satisfaction of students, as well as their development of skills such as: teamworking, communication and problem solving.

## Background

Small group teaching has always been of paramount importance within medical education, with particular emphasis being placed on the benefits of Case Based Learning (CBL) in recent years. CBL has been defined as “a learning and teaching approach that aims to prepare students for clinical practice, through the use of authentic clinical cases. These cases link theory to practice, through the application of knowledge to the cases, and encourage the use of inquiry-based learning methods.” (Thistlethwaite et al., 2021). CBL involves students working within groups to solve clinical and non-clinical cases which enables the development of communication skills and team-working, as well as promoting a deeper understanding of relevant clinical knowledge (McLean, 2016). The development of these skills will then go on to be invaluable within Foundation Years and beyond, creating confident and adaptable doctors (GMC, 2016). Based within a constructivist philosophy (Piaget, 1997; Vygotsky, 1986), CBL encompasses the four basic characteristics of constructivist learning environments (Tam, 2000): 1) knowledge will be shared between teachers and students; 2) teachers and students will share authority; 3) the teacher’s role is one of a facilitator or guide; 4) learning groups will consist of small numbers of heterogeneous students.

Traditionally, CBL sessions are routinely supported by a present, in-person facilitator in a face to face setting, a practice which is both finance and time heavy. A facilitator has been defined as “A person or organization assigned to facilitate progress towards a specific objective, esp. one whose role is to foster communication or understanding within a group of people, or negotiations between various parties; a mediator; a coordinator (esp. of a conference, discussion group, etc.).” (Oxford English Dictionary, 2018). Within the small group pedagogy associated with CBL and Problem Based learning (PBL), the facilitator operationalizes the constructivist philosophy upon which these approaches to learning are based (Salinitri et al., 2015). This is achieved through facilitators fostering a collaborative learning environment in which free discourse and sharing of knowledge in enabled, providing scaffolding to develop and extend students’ knowledge acquisition and problem-solving skills, allowing learners to take ownership of learning process, and challenging learners to consider alternative perspectives and solutions (Salinitri et al., 2015). Facilitating sessions appropriately requires the creation of relevant prompt materials, appropriate faculty training and may not always result in equity of experience across groups. While there is a body of literature that considers the benefits and pedagogy of CBL, including systematic reviews (e.g. Thistlethwaite et al., 2012; McLean, 2016), no evidence synthesis on the role and impact of facilitation could be identified. Among other changes, the recent COVID-19 pandemic has necessitated moves to online learning in many areas (Papapanou et al., 2021), one of which is in the area of CBL. The changes to medical education as a result of the pandemic, expected to be lasting, provide further impetus to consider the role of the facilitator in CBL and digital opportunities to augment facilitation.

This review aims to explore the available literature regarding the benefits of various facilitation approaches (for example: traditional facilitation, staff being available, but not necessarily present within the session as well as self-directed or computer-feedback based direction for students) and to inform conclusions on the advantages and disadvantages of each.

## Aims

To understand the role of facilitation within CBL teaching (in face to face and online settings) and the impact that variation on facilitation styles can have within outcomes such as student experience and engagement.

## Objectives

To conduct a systematic search and review of the available literature on the role and value of facilitation within CBL.

## Methods

### Research questions

- What is the current extent and nature of the research about CBL facilitation in undergraduate medical education?
- What approaches to facilitation of CBL have been used?
- What are the advantages and disadvantages of different approaches to CBL facilitation on a range of outcomes e.g. student experience, student engagement?
- What are the research gaps and limitations of the existing literature?

Following a series of scoping searches to inform the development of a question, a scoping review is suggested for this project to allow for a better understanding of the available literature within this field to be gained. This enables a broad question to be explored which might consider different approaches to facilitation (e.g. in person, online, human or digital) and does not constrain the exploration to considering effects on specific outcomes:

> **Population:** Undergraduate medical students
>
> **Concept:** Approaches to facilitation
>
> **Context:** Case-based learning sessions

Four main academic and medical databases, MedLine, Scopus, Excerpta Medica Database (EMBASE) and Education Research Complete (ERC) will be searched using a variety of search terms and MeSH headings (Table 1):

**Table 1:**
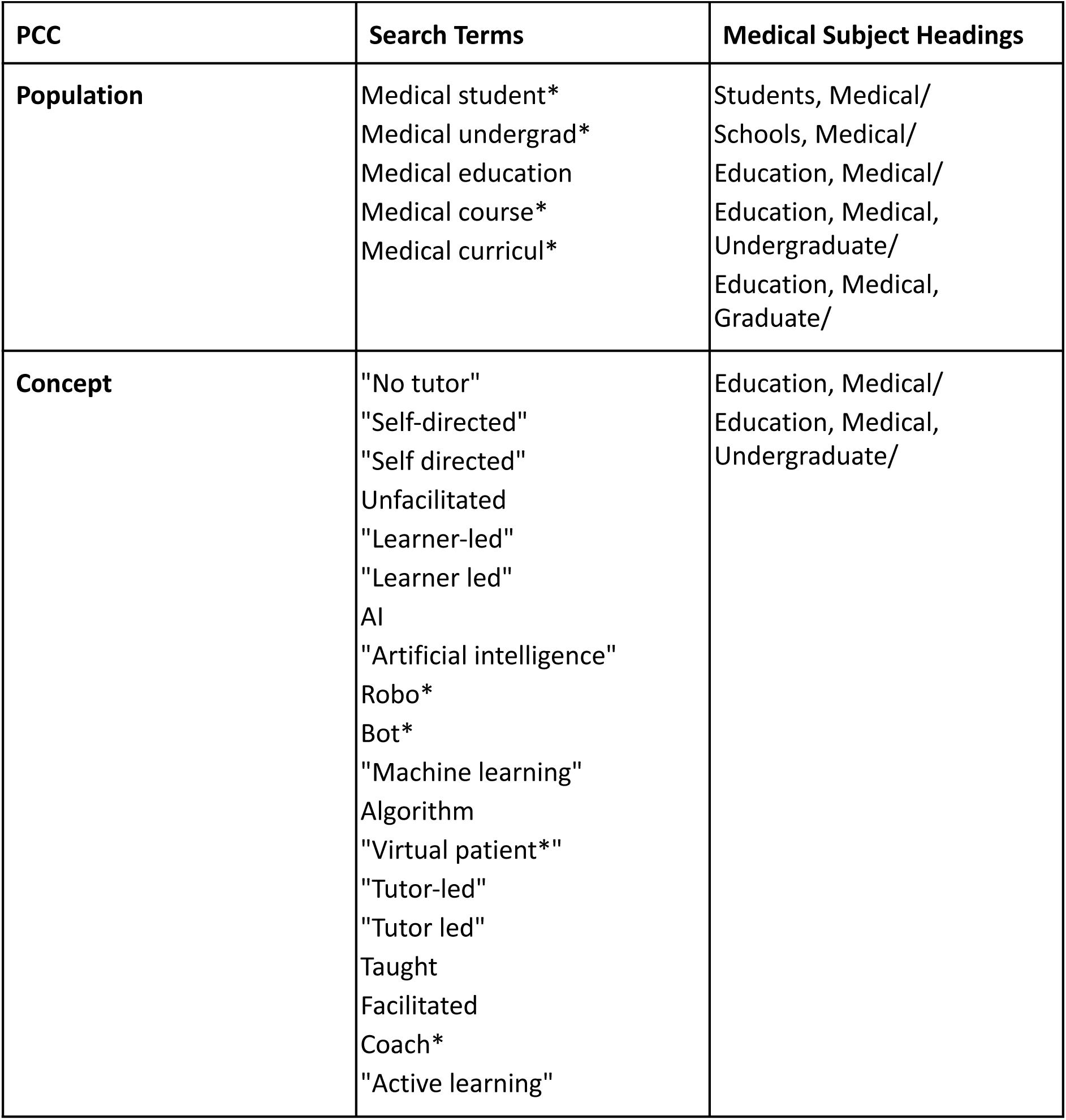
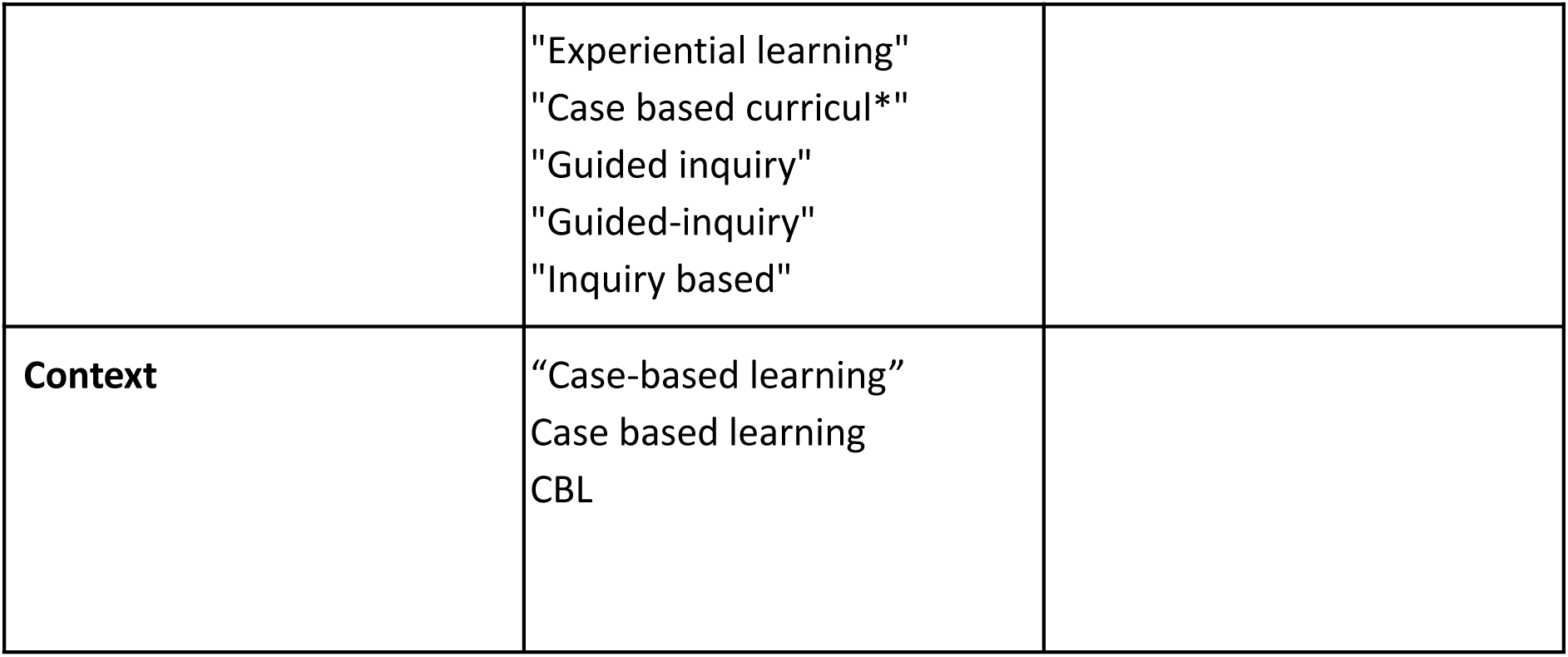
Development of search terms.

The full search strategy for Medline search is located in Appendix 1.

### Study selection

The process outlined in Preferred Reporting Items for Systematic reviews and Meta-Analyses extension for Scoping Reviews (PRISMA-ScR) for the searching, source selection, data charting, and reporting (Tricco *et al*., 2018) will be followed. The search results will be imported into Endnote and all duplicates removed. Covidence (https://www.covidence.org/) will be used to undertake the screening and review process.

Two reviewers will independently screen the titles and abstracts of all articles against the inclusion and exclusion criteria (Table 2). In the case of agreement between both reviewers, the article will be included and the full text version will be sought for further screening. At all stages of the screening process, in the case of disagreement over whether an article should be included, the reviewers will meet to discuss and if a consensus is not reached, a third reviewer will adjudicate. The full texts of the selected articles will be reviewed against the inclusion and exclusion criteria, and a final selection for the synthesis stage will be obtained. The reviewers will meet at the beginning, midpoint and end of the screening process to discuss any problems and review the search strategy if required (Levac *et al*., 2010). In accordance with scoping review methodology, it is anticipated that the selection criteria may be further developed and iterated as the study selection is undertaken.

**Table 2:** Inclusion and exclusion criteria.

| Inclusion Criteria | Exclusion Criteria |
| --- | --- |
| Medical students studying at an undergraduate level.<br>Teaching with a CBL mode of delivery*<br>Peer-reviewed papers not limited by article type | Undergraduate students on non-medical courses and those undertaking postgraduate or specialist training in a medical field.<br>Studies written in a language other than English.<br>Studies focusing on CBL tutor faculty development. |
|  | • Studies which focus on facilitation within teaching other than CBL (e.g. PBL, Team based learning (TBL)).<br>• Studies that identify facilitation as part of the CBL process but do not comment/evaluate it beyond identifying its presence |

\*It became apparent after several scoping searches that, due to the many variations on the CBL model and a broad application of the terminology within the literature, that clear criteria would need to be outlined to ensure comparability within the studies being selected. Therefore, the below criteria have been outlined as being in line with the majority of current CBL practices:

#### Classification of CBL to be used in this study

- Involves a focused, authentic case through which students can holistically consider a patient instead of via separate views of diseased organs and link classroom-based theory to clinical practice through integration of basic science with clinical management (Chan et al. 2008)
- Students work in small groups to collaboratively direct their own learning towards defined learning outcomes.
- Involves a case that extends over a time period of more than one scheduled session.

### Data charting

In accordance with scoping review terminology, data extraction will be referred to as ‘data charting’ (Peters *et al*., 2020). A data charting form will be developed and piloted (Table 3). As data charting is an iterative process, it is anticipated that the data charting form may be further updated during the data extraction process. Two reviewers will independently chart the data with any disagreements will be resolved by discussion and consensus. In the event that a consensus cannot be reached, a third reviewer will adjudicate. Charting will take place in Covidence.

**Table 3:** Charting Form.

| Evidence source details and characteristics |  |
| --- | --- |
| Citation details | Author/s, date, title, journal, volume, issue, pages |
| Country in which the study was conducted |  |
| Aims and purpose | As stated by authors |
| Research approach | E.g. details of study design, data collection methods |
| Approach to facilitation | Description of approach to facilitation e.g. no facilitation, tutor, digital |
| Findings/commentary about facilitation | E.g., results of process, outcomes of intervention, key points in discussion, conclusions drawn by authors |
| Conclusions/recommendations regarding facilitation | As stated by authors |

### Quality assessment

Dependent on the types of studies that the search identifies, a critical appraisal/quality assessment tool may be used. This will be determined when the study selection is complete and the types of included evidence are known.

### Data synthesis

A descriptive numerical summary will be produced, including reporting of the research approaches used and the study findings/conclusions (Arksey & O’Malley, 2005; Levac *et al*., 2010). The frequency of research approaches employed and approaches to facilitation described will also be analysed and presented (Peters *et al*., 2020). The aim of this will be to map the data and identify gaps in the research. The authors will then use a narrative synthesis approach to summarise and explain the main themes that emerge from the studies, linked to the research questions and conceptual framework (Levac *et al*., 2010). As CBL is rooted in the constructivist philosophy of learning where knowledge is constructed by learners through their interaction with the environment and from their life experiences (Piaget, 1997; Vygotsky, 1986) and encompasses the four basic characteristics of constructivist learning environments (Tam, 2000): 1) knowledge will be shared between teachers and students; 2) teachers and students will share authority; 3) the teacher’s role is one of a facilitator or guide; 4) learning groups will consist of small numbers of heterogeneous students, a constructivist conceptual framework will be used to guide the synthesis. This will be an iterative process and will depend on the studies and data uncovered in the search. The aim will be to produce a descriptive account of the literature, addressing the current evidence for different approaches to facilitation of CBL, the research approaches being undertaken, and areas for future research and development.

## Conclusions

Overall, it is hoped that this review will be able to comment on the role of facilitation within CBL sessions and the potential impact that this has on engagement and satisfaction of students, as well as their development of skills such as: teamworking, communication, problem solving and attainment. This information will therefore be valuable for indicating potential areas for further research within the field of CBL facilitation.

## Data Availability

All data referred to within the manuscript is available within public databases online.

## Appendix 1- Medline search strategy

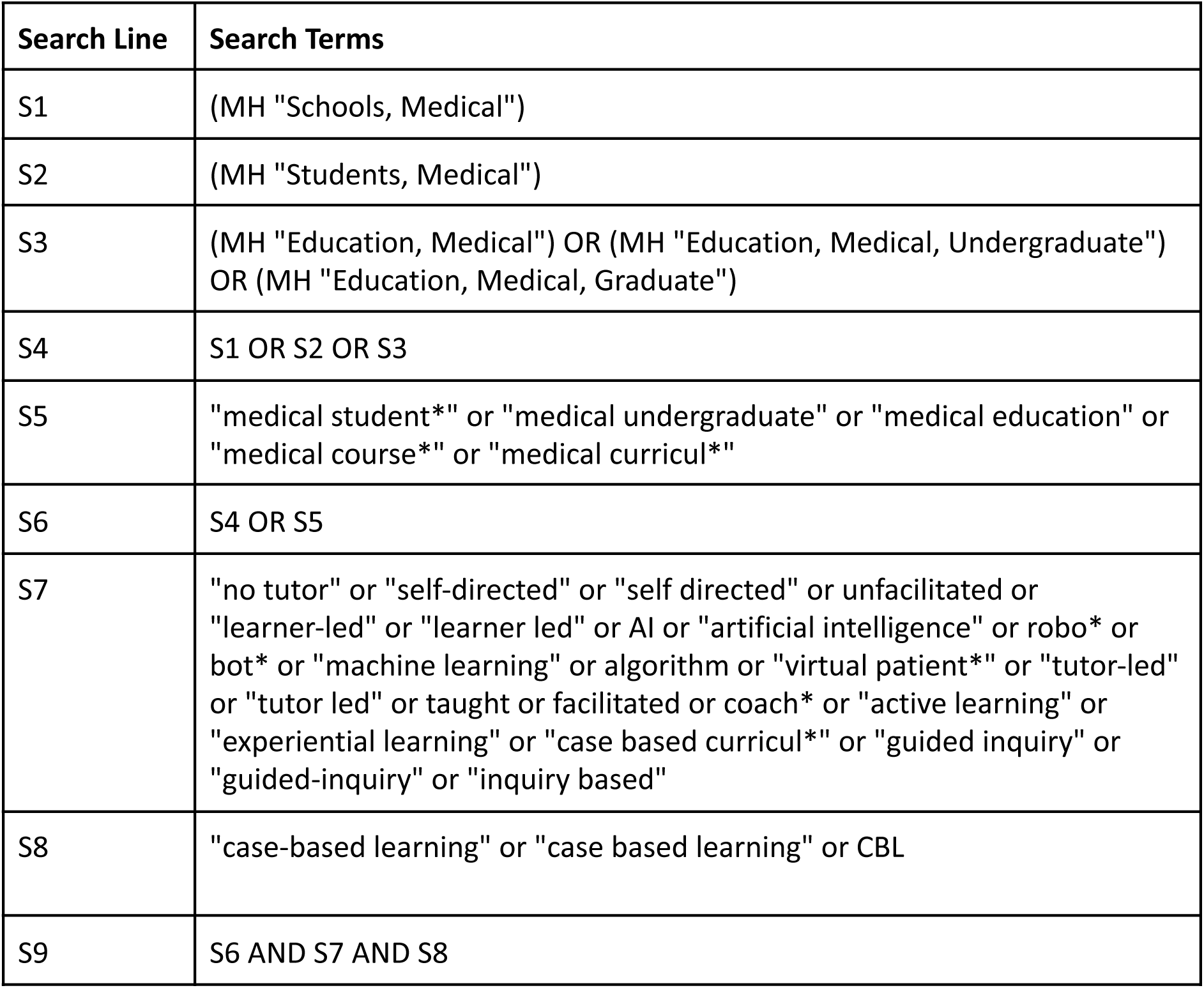

Appendix 1 - Medline search strategy
| Search Line | Search Terms |
| --- | --- |
| S1 | (MH "Schools, Medical") |
| S2 | (MH "Students, Medical") |
| S3 | (MH "Education, Medical") OR (MH "Education, Medical, Undergraduate")<br>OR (MH "Education, Medical, Graduate") |
| S4 | S1 OR S2 OR S3 |
| S5 | "medical student*" or "medical undergraduate" or "medical education" or<br>"medical course*" or "medical curricul*" |
| S6 | S4 OR S5 |
| S7 | "no tutor" or "self-directed" or "self directed" or unfacilitated or<br>"learner-led" or "learner led" or AI or "artificial intelligence" or robo* or<br>bot* or "machine learning" or algorithm or "virtual patient*" or "tutor-led"<br>or "tutor led" or taught or facilitated or coach* or "active learning" or<br>"experiential learning" or "case based curricul*" or "guided inquiry" or<br>"guided-inquiry" or "inquiry based" |
| S8 | "case-based learning" or "case based learning" or CBL |
| S9 | S6 AND S7 AND S8 |

## References

Arksey, H. & O’Malley, L. (2005) Scoping studies: towards a methodological framework. International Journal of Social Research Methodology, 8 (1): 19–32.

Chan WP, Hsu CY, Hong CY. 2008. Innovative ‘‘Case-Based IntegratedTeaching’’ in an undergraduate medical curriculum: Development and teachers’ and students’ responses. Ann Acad Med Singapore, 37, (11):952–956.

General Medical Council, 2016. Achieving Good Medical Practice. London: General Medical Council.

Levac, D., Colquhoun, H. & O’Brien, K. K. (2010) Scoping studies: advancing the methodology. Implementation Science, 5, article no: 69. Available from: https://doi.org/10.1186/1748-5908-5-69 (xAccessed 15/03/2021).

McLean, S., 2016. Case-Based Learning and its Application in Medical and Health-Care Fields: A Review of Worldwide Literature. Journal of Medical Education and Curricular Development, [online] 3, p.JMECD.S20377. Available at: <https://www.ncbi.nlm.nih.gov/pmc/articles/PMC5736264/> [Accessed 5 August 2021]

Oxford English Dictionary (2018) https://www.oed.com/

Peters, M. D. J., Godfrey, C., McInerney, P., Munn, Z., Tricco, A. C. & Khalil, H. (2020) Chapter 11: Scoping Reviews (2020 version). In: Aromataris, E. & Munn, Z. eds. JBI Manual for Evidence Synthesis. JBI. Available from: https://synthesismanual.jbi.global (Accessed 14/03/2021).

Papapanou, M., Routsi, E., Tsamakis, K., et al. (2021). Medical education challenges and innovations during COVID-19 pandemicPostgraduate Medical Journal. doi: 10.1136/postgradmedj-2021-140032

Piaget, J. (1997) The moral judgment of the child. New York, NY: Simon and Schuster.

Salinitri, F. D., Wilhelm, S. M., & Crabtree, B. L. (2015). Facilitating Facilitators: Enhancing PBL through a Structured Facilitator Development Program. Interdisciplinary Journal of Problem-Based Learning, 9(1). Available at: https://doi.org/10.7771/1541-5015.1509

Tam, M. (2000). Constructivism, Instructional Design, and Technology: Implications for Transforming Distance Learning. Educational Technology and Society, 3 (2), 50–60.

Thistlethwaite, J., Davies, D., Ekeocha, S., Kidd, J., MacDougall, C., Matthews, P., Purkis, J. & Clay, D. (2012). The effectiveness of case-based learning in health professional education. A BEME systematic review: BEME Guide No. 23,Medical Teacher, 34:6, e421–e444, DOI: 10.3109/0142159X.2012.680939

Tricco, A. C., Lillie, E., Zarin, W., O’Brien, K. K., Colquhoun, H., Levac, D., Moher, D., Peters, M. D., Horsley, T., Weeks, L., Hempel, S. et al. (2018) PRISMA extension for scoping reviews (PRISMA-ScR): checklist and explanation. Annals of Internal Medicine, 169 (7): 467–473.

Vygotsky, L. (1986). Thought and language. Cambridge, MA:MIT Press.

